# DXA-derived hip shape is associated with hip fracture: a longitudinal study of 38,123 UK Biobank participants

**DOI:** 10.1101/2025.09.30.25336960

**Authors:** Sophie Scott, Asad Hashmi, Raja Ebsim, Fiona R Saunders, Jennifer S Gregory, Richard M Aspden, Claudia Lindner, Timothy Cootes, Nicholas C Harvey, Jonathan H Tobias, Benjamin G Faber, Rhona A Beynon

## Abstract

Despite advancements in fracture prediction tools and osteoporosis management, hip fractures remain a significant consequence of bone fragility, with a 22% one-year mortality. Hip geometric measures (GMs) have been associated with fracture risk; however, their strong correlation hinders the identification of independent influences, leaving their relative predictive value unclear. Statistical shape modelling (SSM) provides a more holistic assessment of hip shape compared to using pre-determined GMs. This study aimed to evaluate whether SSM-derived hip shape from dual-energy X-ray absorptiometry (DXA) scans can predict hip fracture, independently of individual GMs. Previously, we applied SSM to left hip DXA images in UK Biobank, a large prospective cohort with linked hospital records, generating ten orthogonal hip shape modes (HSMs), that explained 86% of shape variance. Additionally, femoral neck width (FNW), femoral head diameter (FHD), and hip axis length (HAL) were derived from these DXAs. In the current analysis, Cox proportional hazard models, adjusted for age, sex, height, weight, bone mineral density (BMD), and GMs (FNW, HAL, FHD), were used to examine the longitudinal associations between each HSM and first incident hospital diagnosed hip fracture. A Bonferroni adjusted p-value threshold (p<0.004) was used to account for the 13 exposures. Among the 38,123 participants (mean age 63.7 years; 52% female; mean follow-up 5 years), 133 (0.35%) experienced subsequent hip fracture. HSM2, characterised by a narrower FNW, a higher neck shaft angle, and reduced acetabular coverage, showed a strong association with hip fracture risk (HR 1.32, 95% CI 1.11-1.58, P 1.47×10^-3^), which persisted after full adjustment (1.30, 1.09-1.55, 3.27×10^-3^). There was no evidence for an association with other HSMs. These findings suggest that DXA-derived hip shape is associated with hip fracture risk independently of BMD and GMs. Incorporating global hip shape into fracture risk assessment tools could enhance prediction accuracy and inform targeted interventions.

**Lay summary:** Despite improvements in hip fracture prevention, they remain a major problem, with 22% of people dying within a year of sustaining one. This study looked at medical images from 38,123 individuals in UK Biobank to assess the shape of their hip using computer-aided statistical techniques. The results indicate that a hip shape variation describing a narrower femoral neck and a larger angle linking the neck and the femoral shaft is linked to fracture. This association persisted after accounting for other known hip shape measures related to fracture risk. Therefore, hip shape could help improve prediction and prevention of hip fractures.

## Introduction

The annual number of hip fractures in the UK is projected to rise by 32% over the next 4 years^1^, highlighting the need for accurate prediction of hip fracture risk. These fractures represent a significant consequence of osteoporosis-related bone fragility and carry a one-year mortality rate of 22%^2^. However, not all individuals who sustain a hip fracture meet the diagnostic criteria for osteoporosis^3^, which is primarily based on bone mineral density (BMD). Clinical risk assessment tools such as FRAX^®4^ – widely used in over 100 international guidelines – and the UK-specific Qfracture^5^, have been developed to better predict the risk of incident fractures, but still lack optimal sensitivity^6,7^. Consequently, incorporating additional factors not currently considered in existing tools could help improve the accuracy of fracture risk prediction^8^.

Variation in hip shape is increasingly recognised as a contributor to hip fracture^9,10^, having also been linked to osteoarthritis^11^. Hip shape can be assessed through measuring individual geometric measures (GMs), or by evaluating the overall shape. Common examples of GMs include hip axis length (HAL), neck shaft angle (NSA), femoral neck width (FNW), and femoral head diameter (FHD), which can all be derived from DXA scans, either manually or using software such as Hip Structural Analysis^12^. Although evidence linking GMs to fracture risk is inconsistent, a recent meta-analysis found that increased HAL, NSA, and FNW are associated with higher fracture risk, with pooled odds ratios (OR) of 1.53, 1.47, and 2.68 respectively^10^. This did not account for factors such as age and sex. Nonetheless, the International Society of Clinical Densitometry recommends using only HAL for assessing hip fracture risk in females, and advises against using GMs to guide treatment decisions^12^. Moreover, the high correlation between GMs such as FNW and HAL^11^, as well as the correlation between GMs and body size^13^, complicates the evaluation of their individual contributions to hip fracture risk. In contrast, assessing hip shape as a whole, rather than focusing on individual GMs, may provide a more comprehensive understanding of hip health and fracture risk by accounting for overall morphology and the relationships between different features^14^.

Statistical shape modelling (SSM), a computer-aided technique designed to capture the statistical variability of shapes within a dataset^15^, can be used to provide a more holistic measure of hip shape. SSM uses outline points derived from hip images and employs principal component analysis (PCA) to produce orthogonal modes of shape variation, termed hip shape modes (HSM)^16^, which each capture a different aspect of hip morphology. Although research linking HSMs to hip fracture risk is limited, one study that applied SSM to radiographs found that a HSM characterised by a longer femoral neck, smaller femoral head and a narrower FNW was associated with a higher fracture risk (OR 2.48)^8^. Studies comparing SSM-derived measures of hip shape to GMs in the context of hip fractures have been limited to small studies^9^, which have been unable to show that SSM-derived hip fracture risk is independent of GMs. This underscores the need for a comparative analysis to identify the most effective predictors of hip fracture risk. In our recent work using UK Biobank (UKB) we developed a machine-learning algorithm that automatically places outline points on high resolution hip DXA images^17^, facilitating the generation of hip shape measures in large numbers.

In the present study, we aimed to establish whether SSM-derived hip shape, obtained using our automated point placement method, is associated with hip fracture risk independently of established risk factors and hip GMs, while also analysing potential sex differences within these associations in the UKB cohort.

## Methods and materials

### Population

UKB is a prospective cohort study that recruited ∼500,000 males and females, aged 40-69 years, from 22 assessment centres across the UK between 2006-2010^18^. Baseline genetic and phenotypic information was obtained through questionnaires, physical measurements and biological samples. In 2014, UKB launched the Image Enhancement study, which aims to gather imaging data, including hip DXA scans, from 100,000 participants^19^. For this study, ∼40,000 left-hip DXA images with outline points delineating the bone contour were available (October 2023). This study is overseen by the UKB Ethics Advisory Committee, and ethical approval was given by the National Information Governing Board for Health and Social Care and North-West Multi-centre Research Ethics committee (11/NW/0382). All participants provided informed consent for their data to be used in the study.

### Acquisition of DXA-derived hip shape

Hip DXA images were acquired following a standardised protocol using an iDXA scanner (GE-Lunar, Madison, WI, USA), with participants’ legs positioned at an internal rotation of 15-25°^19^. A Random Forest-based machine learning algorithm^20^ (BoneFinder®, The University of Manchester) had been previously used to automatically place the hip outline points^17^. This algorithm was initially trained on a subset of ∼7,000 manually marked-up images before being applied to the remaining ∼33,000 images. A total of 85 outline points were placed around the femoral head and acetabulum, including the greater and lesser trochanters (Figure 1). The placement of the outline points was manually verified, with only 10% requiring adjustment and an average correction distance of 1.9 mm.

**Figure 1:**
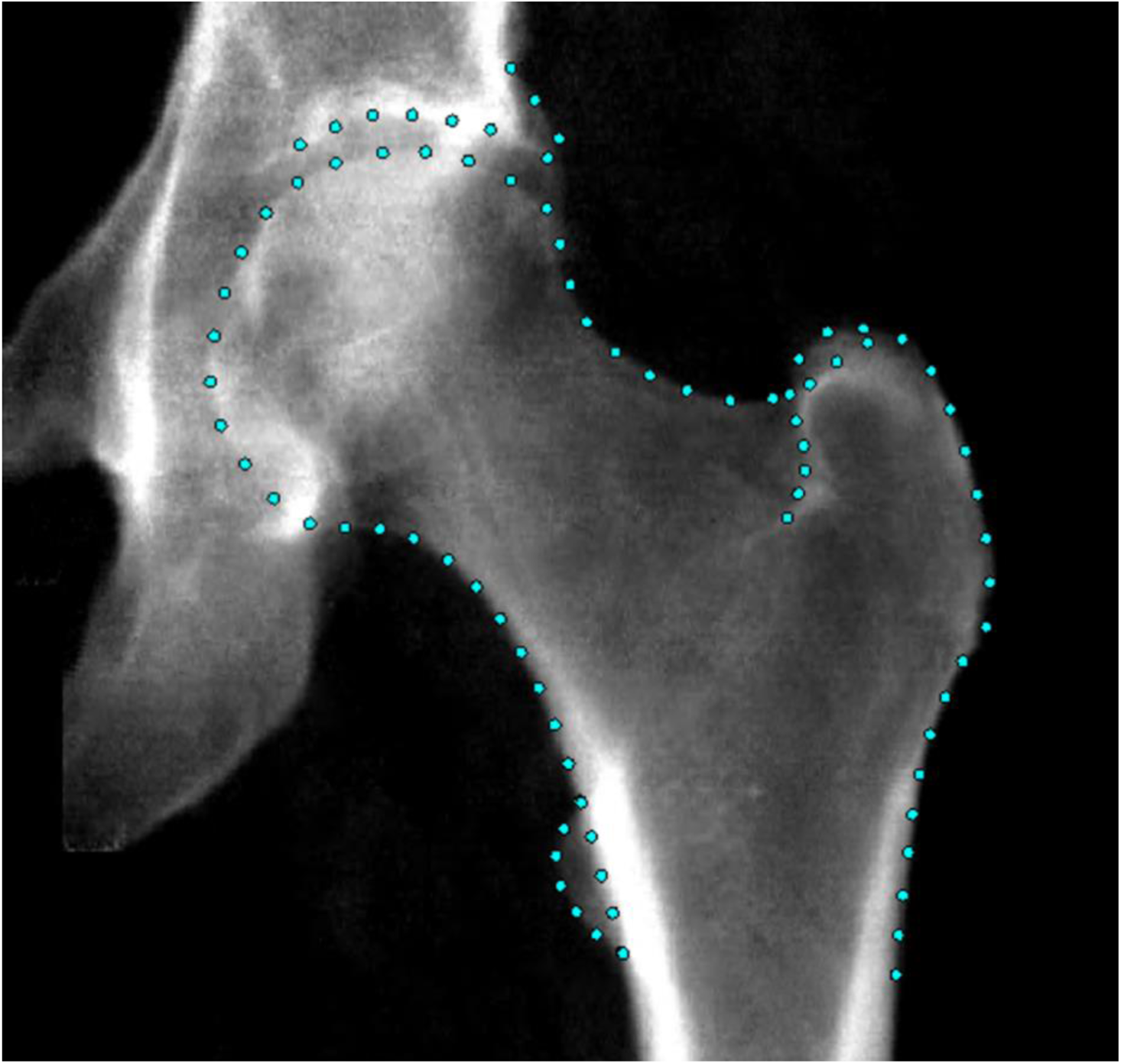
An example hip DXA scan from UKB showing the points placed around the hip joint.

Once outlined, principal component analysis (PCA) was performed to generate a set of orthogonal hip shape modes (HSMs), which collectively explain 100% of the variance^17^. To minimise the burden of multiple testing, this analysis focused on the first ten HSMs, which accounted for 86% of the shape variance within the data set. Subsequent HSMs explained minimal additional shape variance and are unlikely to hold clinical significance. Additionally, FNW, FHD, and HAL were previously derived from the DXA scans using an openly available custom Python script, as described elsewhere^11,21^.

### Ascertainment of hip fracture

Hip fracture data were obtained through linkage to hospital episode statistics (HES), which uses the International Classification of Diseases (ICD) 10^th^ revision codes. Hip fractures were identified based on the following codes: fractured neck of femur (S72.0), pertrochanteric fracture (S72.1), subtrochanteric fracture (S72.2), stress fracture, not elsewhere classified (Pelvic region and thigh) (M84.359), or pathological fracture, not elsewhere classified (Pelvic region and thigh) (M84.459). Recording of HES data began on the 1^st^ April 1997. Hip fracture data were downloaded in August 2023, capturing information up until the end of October 2022.

### Statistical analysis

Descriptive statistics, including means, standard deviations (SDs), and ranges, were used to summarise population characteristics and the distributions of HSMs and GMs. Histograms were plotted for each HSM to confirm normal distribution. The correlation between each HSM, GM, BMD and demographic factors (height, weight, age), were assessed using Pearson correlation co-efficient (r). Cox proportional hazard models were used to examine the longitudinal associations between each HSM and hip fracture risk, as well as between each GM (FNW, FHD, HAL), and hip fracture risk. The follow-up period concluded at the earliest event, which was either the first incident hip fracture during follow-up, withdrawal, censoring due to death or until the end of the study (31/10/2022). Individuals who had a hip fracture before attending the imaging assessment, i.e. before the DXA scan, were excluded from the analysis. The Cox proportional hazards assumption was tested using the Schoenfeld residuals approach. A Bonferroni adjusted p-value threshold (*P*<0.004) was used to account for the thirteen exposures tested (ten HSMs and three GMs). Results are shown as hazard ratios (HR), which represent the relative risk of experiencing a hip fracture over time, with 95% CI and p-values.

Results are presented across four models: Model 1 is unadjusted; Model 2 adjusts for demographic characteristics (age, sex, height, and weight); Model 3 additionally adjusts for left hip femoral BMD; and Model 4 further adjusts for GMs (FNW, FHD, and HAL). When a GM is the exposure, model 4 adjusts for the other two GMs. Both combined-sex and sex-stratified analyses were conducted to account for known disparities in fracture risk^22^ and hip shape^23^ between males and females. All statistical analyses were performed using STATA version 18 (Stata Corp, College Station, TX, USA).

### Composite models

To investigate the overall at-risk hip shape for fracture a composite HSM figure was plotted by combining all HSMs. Briefly, to do this, unadjusted beta coefficients for the associations between HSMs and fracture were first computed. Each beta was then multiplied by 10 to enhance the visualisation of shapes, and subsequently multiplied by the HSM-specific SD to account for the contribution of each HSM to the overall shape variance. These adjusted values were combined into a single vector to assess the collective impact of hip shape on hip fracture.

## Results

### Baseline characteristics

This study included 38,128 UKB individuals with complete data and a left hip DXA image available (Table 1). The mean age was 63.7 years, and 52% of participants were female. Mean BMD of the left femur was 0.99 g/cm^2^, with females having a lower mean BMD (0.93 g/cm^2^) compared to males (1.06 g/cm^2^). A total of 133 participants (0.35%) had a hip fracture, with a higher prevalence among females (89 cases, 0.45%) compared with males (44 cases, 0.24%). HSMs 1-10 had a mean value of 0 by design (Figure 2). Mean HSM values differed between sexes, with the greatest difference seen in HSM1, HSM3, and HSM9. For the GMs, the combined sex mean for FNW was 31.6 mm, FHD was 45.9 mm and HAL was 96.7 mm. Males had a greater mean FNW, FHD, and HAL than females.

**Table 1.** Descriptive statistics of UK Biobank participants included in this study.

|  | **Combined** | **Female** | **Male** |
| --- | --- | --- | --- |
|  | N = 38,123 | N = 19,820 (52%) | N = 18,303 (48%) |
| **Exposures** | **Mean [SD, Range]** | **Mean [SD, Range]** | **Mean [SD, Range]** |
| Age (years) | 63.7 [7.6, 44-82] | 63.0 [7.4, 45-82] | 64.3 [7.7, 44-81] |
| Height (cm) | 170.2 [9.4, 135-204] | 163.7 [6.4, 135-196] | 177.2 [6.6, 150-204] |
| Weight (kg) | 75.4 [15.1, 34-171] | 68.2 [12.9, 34-169] | 83.2 [13.4, 47-171] |
| Left femoral bone mineral density (g/cm<sup>2</sup>) | 1.0 [0.2, 0.0-1.7] | 0.9 [0.1, 0.1-1.7] | 1.1 [0.1, 0.0-1.7] |
| Hip shape mode 1 | 0.0 [1.0, -4.6-3.9] | 0.3 [0.9, -3.8-3.9] | -0.3 [1.0, -4.6-3.6] |
| Hip shape mode 2 | 0.0 [1.0, -4.7-4.5] | -0.0 [1.0, -4.7-4.2] | 0.0 [1.0, -4.5-4.5] |
| Hip shape mode 3 | 0.0 [1.0, -4.1-4.3] | -0.3 [0.9, -4.1-4.0] | 0.3 [1.0, -3.6-4.3] |
| Hip shape mode 4 | 0.0 [1.0, -4.4-4.0] | -0.1 [1.0, -4.4-4.0] | 0.1 [1.0, -3.8-4.0] |
| Hip shape mode 5 | 0.0 [1.0, -4.5-3.5] | 0.0 [1.1, -4.4-3.4] | -0.0 [0.9, -4.5-3.5] |
| Hip shape mode 6 | 0.0 [1.0, -4.6-5.0] | 0.2 [1.0, -3.4-5.0] | -0.2 [1.0, -4.6-3.9] |
| Hip shape mode 7 | 0.0 [1.0, -4.9-5.0] | 0.1 [1.0, -4.9-4.7] | -0.1 [1.0, -4.6-5.0] |
| Hip shape mode 8 | 0.0 [1.0, -4.4-4.5] | -0.1 [1.0, -4.4-4.0] | 0.1 [1.0, -4.0-4.5] |
| Hip shape mode 9 | 0.0 [1.0, -4.1-5.0] | -0.3 [0.9, -4.1-4.5] | 0.3 [1.0, -3.7-5.0] |
| Hip shape mode 10 | 0.0 [1.0, -4.1-3.8] | 0.0 [1.0, -4.1-3.8] | -0.0 [1.0, -4.1-3.8] |
| Narrowest neck width (mm) | 31.6 [3.5, 21.4-45.8] | 29.0 [2.0, 21.4-37.8] | 34.5 [2.4, 22.9-45.8] |
| Diameter of femoral head (mm) | 45.9 [3.8, 33.4-64.4] | 43.0 [2.3, 33.4-53.7] | 49.0 [2.6, 34.7-64.4] |
| Hip axis length (mm) | 96.7 [8.0, 68.1-127.1] | 90.8 [4.8, 68.1-115.5] | 103.1 [5.5, 76.9-127.1] |
|  | **Number fractured [%]** | **Number fractured [%]** | **Number fractured [%]** |
| Hospital diagnosed fracture | 133 [0.35] | 89 [0.45] | 44 [0.24] |
|  | **Mean [SD, Range]** | **Mean [SD, Range]** | **Mean [SD, Range]** |
| Time to end of study (years) | 5.0 [1.5, 0.2-8.5] | 5.0 [1.5, 0.1-8.5] | 5.0 [1.5, 0.2-8.5] |

Population characteristics of the UK Biobank participants included in this study with complete data for all covariates.

**Figure 2:**
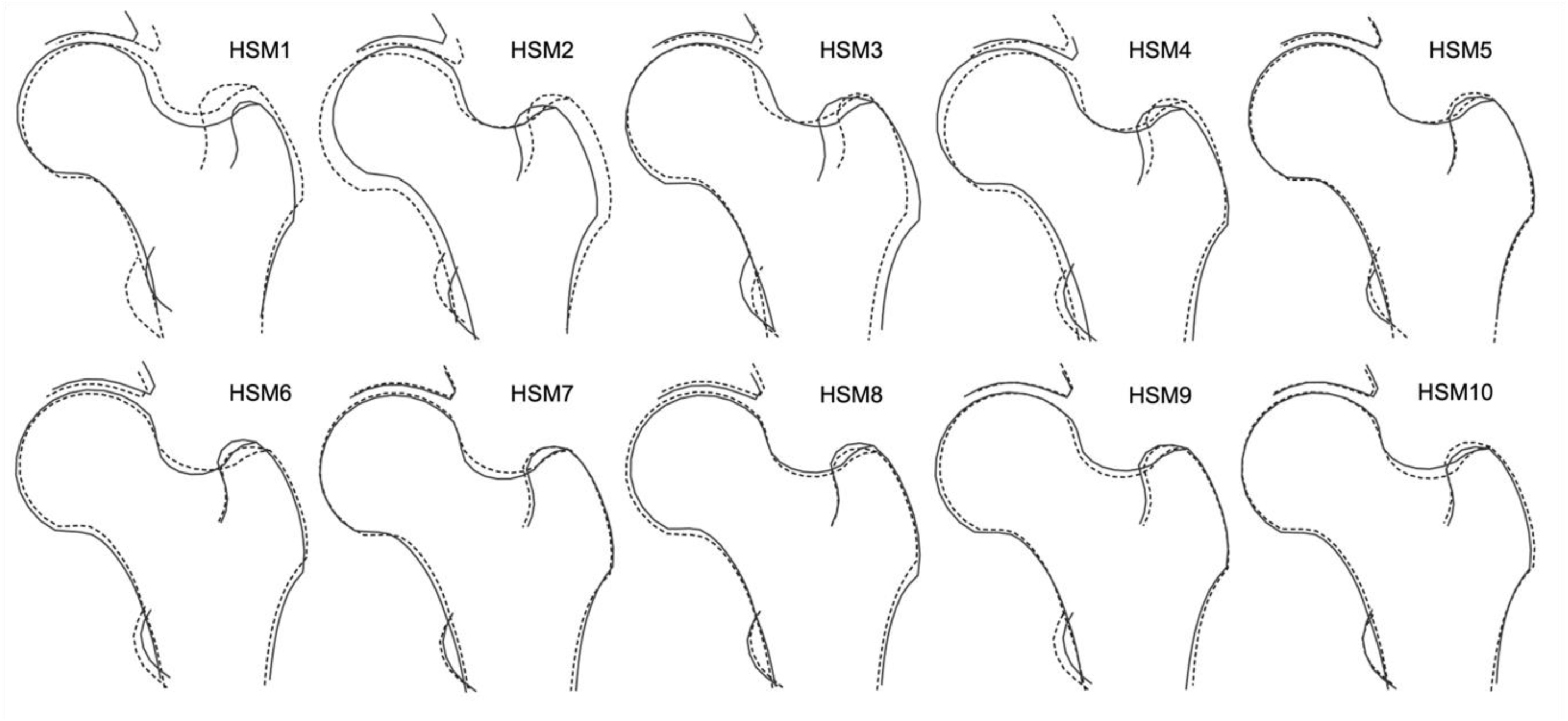
The ten hip shape modes (HSMs). The solid line shows the shape +2 standard deviations (SD) from the mean, and the dotted line shows the shape −2 SDs from the mean.

**Figure 3:**
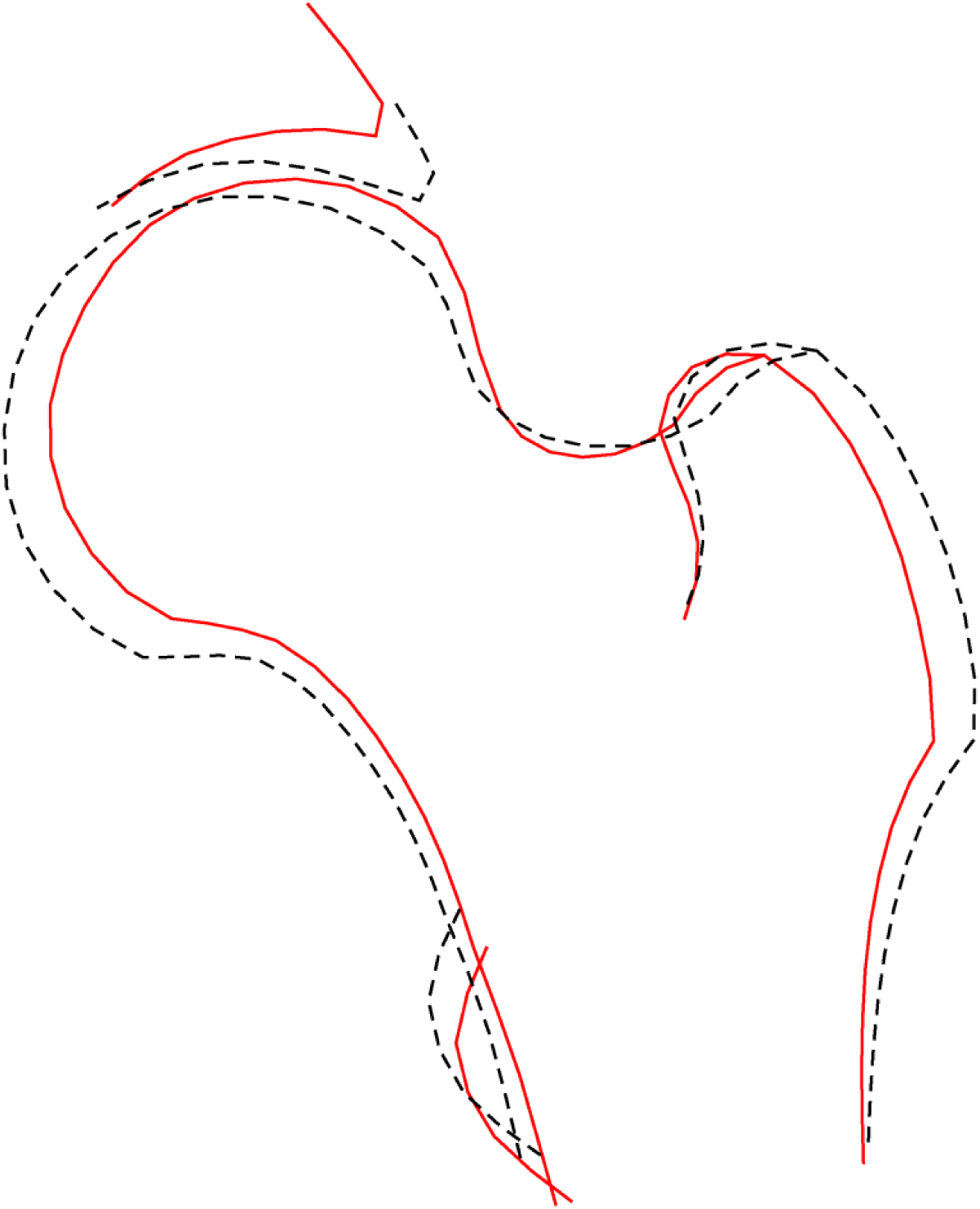
Composite image of the ten hip shape modes. The solid line shows the shape at risk of fracture, the dotted line shows the mean shape.

**Figure 4:**
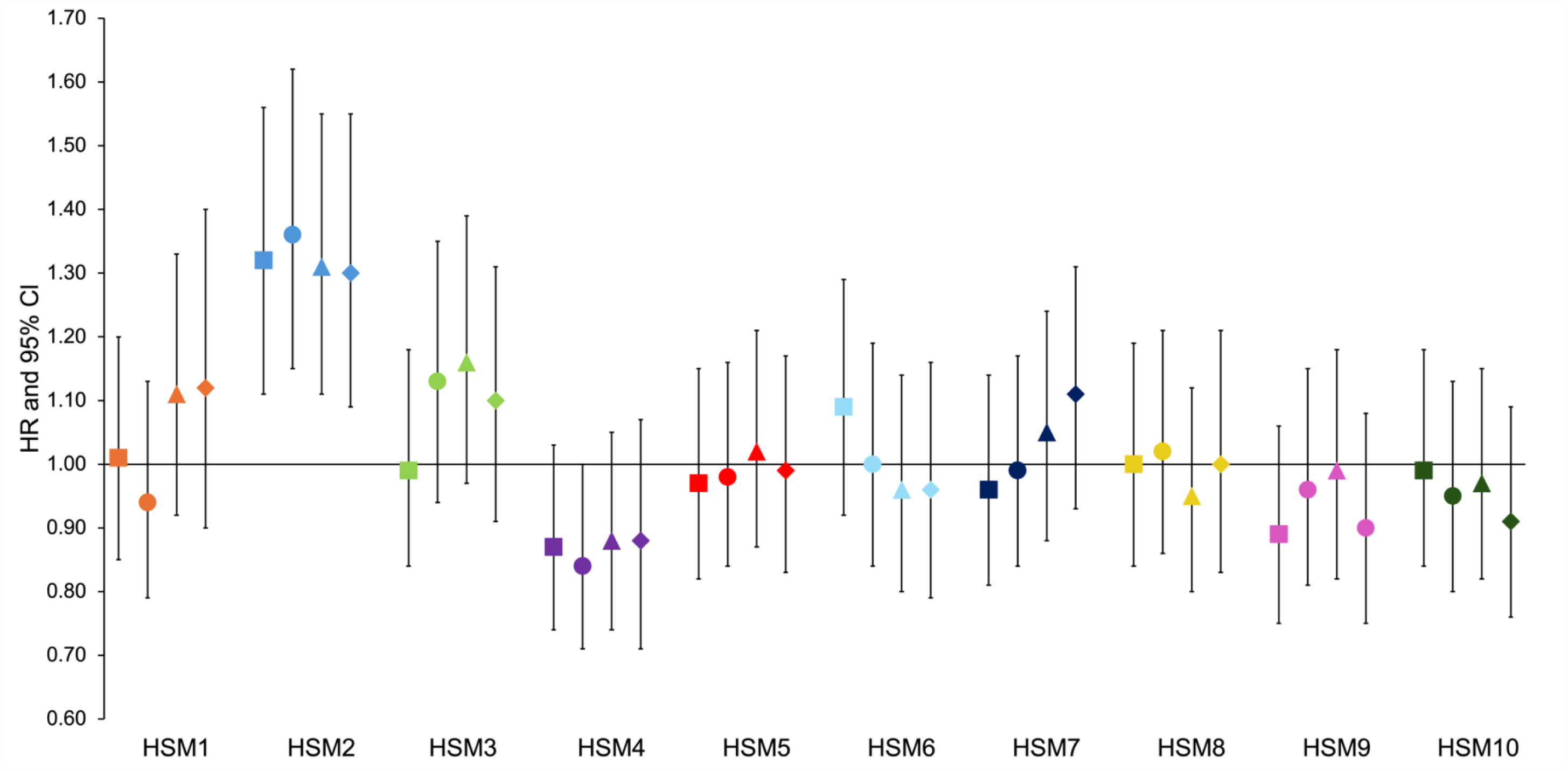
Cox proportional hazard results for the association between each hip shape mode (HSM) and hip fracture in combined sex analysis. Hazard ratios (HR) with 95% confidence intervals (CI) are plotted. Square = unadjusted (model 1); circle = adjusted for age, sex, height, and weight (model 2); triangle = adjusted for model 2 plus bone mineral density (model 3); diamond = fully adjusted for model 3 plus the three geometric measures (model 4).

### Geometric measures and their inter-relationships

FHD, FNW, and HAL were all highly correlated with each other (r 0.81-0.89) and with height (r 0.75-0.81) (Supplementary Figure 1). Weight was moderately correlated with FHD, FNW, HAL, height, and BMD (r 0.52-0.57). The HSMs were orthogonal by design. Similarly, no correlation was observed between the HSMs and the other covariates.

### Geometric measures and their association to hip fracture

#### Femoral Neck Width

In the unadjusted analysis of all participants (Figure 5, Supplementary Table 2), FNW was not associated with hip fracture (Model 1: 1.15, 0.97-1.36, 0.11). However, a strong association was seen between a wider FNW and hip fracture following adjustment for demographic characteristics and BMD (Model 3: 1.77, 1.30-2.43, 3.26×10^-4^). In sex-stratified analysis (Supplementary Table 2), a wider FNW showed a strong association with hip fracture in both sexes, both in the unadjusted model and following adjustment for demographic characteristics. In males, the strongest association was observed in the unadjusted model (Model 1: 2.17, 1.44-3.25, 1.99×10^-4^). The association weakened with adjustment for BMD (Model 3: 1.75, 1,08-2.82, 0.02). A similar trend was noted in females, with the strongest association being in the unadjusted model (Model 1: 2.88, 2.05-4.06, 1.40×10^-9^). Further adjustment for BMD resulted in attenuation (Model 3: 1.70, 1.11-2.59, 0.01). The effect sizes were greater in females compared to males in models 1 and 2, with a similar effect size seen in both sexes in model 3.

**Figure 5:**
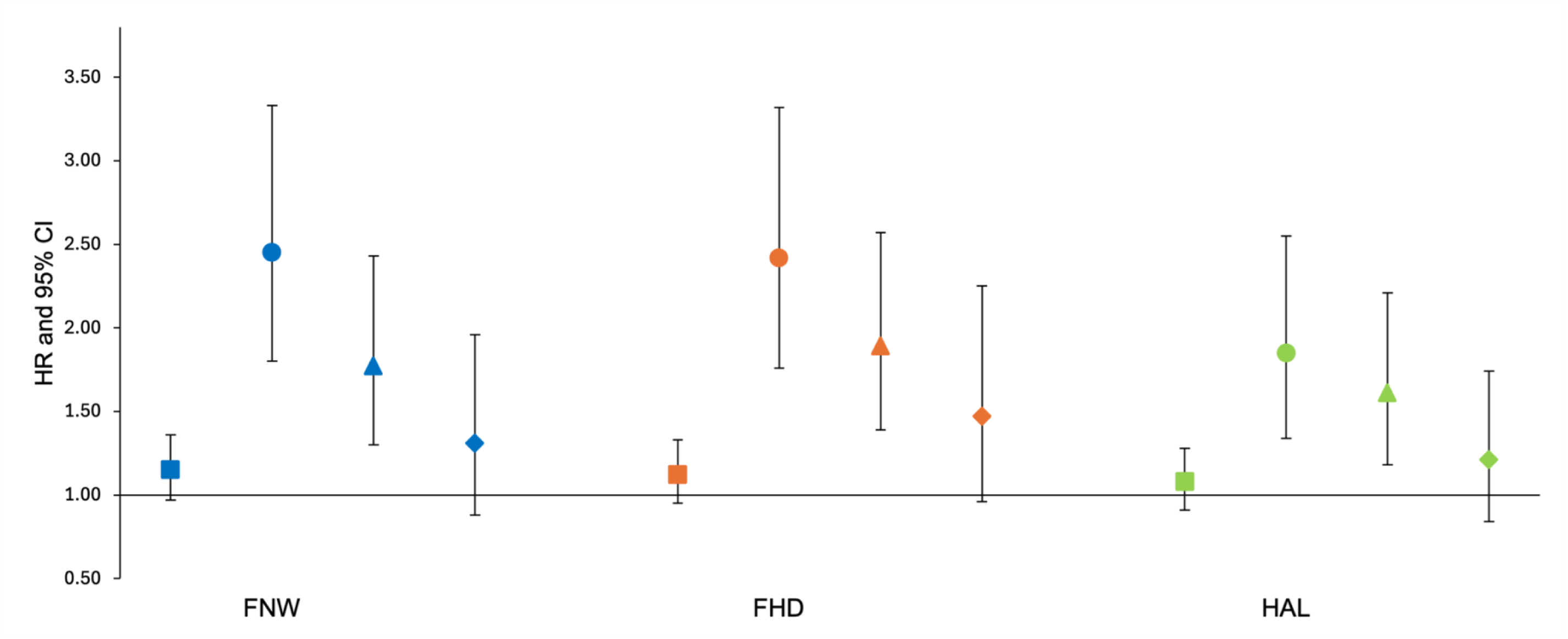
Cox proportional hazard results for the association between each geometric measure (GM) and hip fracture in combined sex analysis. Hazard ratios (HR) with 95% confidence intervals (CI) are plotted. Square = unadjusted (model 1); circle = adjusted for age, sex, height, and weight (model 2); triangle = adjusted for model 2 plus bone mineral density (model 3); diamond = fully adjusted for model 3 plus the other two geometric measures (model 4). FNW = femoral neck width, FHD = femoral head diameter, HAL = hip axis length

### Femoral Head Diameter

In the unadjusted analysis of all participants, there was little evidence for an observed association between FHD and hip fracture (Model 1: 1.12, 0.95-1.53, 0.17) (Figure 5, Supplementary Table 2). However, a strong positive association was seen when adjusting for BMD (Model 3: 1.89, 1.39-2.57, 4.48×10^-5^). In the unadjusted sex-stratified analysis, a larger FHD demonstrated a greater effect size in females compared with males (Model 1: females - 2.43, 1.73-4.30, 2.60×10^-7^; males - 2.30, 1.54-3.44, 4.50×10^-5^). When adjusting for BMD, a larger effect size was seen in males compared with females (Model 3: males - 2.01, 1.28-3.14, 2.26×10^-3^; females - 1.70, 1.11-2.60, 0.01).

### Hip Axis Length

Similar to FNW and FHD, HAL did not show an association with hip fracture in unadjusted analysis of all participants (Model 1: 1.08, 0.91-1.28, 0.39) (Figure 5, Supplementary Table 2). However, an increased HAL was associated with hip fracture after adjusting for BMD (Model 3: 1.61, 1.18-2.21, 3.08×10^-3^). When compared with FNW and FHD, HAL exhibited the smallest effect size across all models. A strong positive association was seen only in the unadjusted sex-stratified analysis (Model 1: males - 2.07, 1.37-3.11, 4.84×10^-4^; females - 2.05, 1.48-2.85, 1.58×10^-5^) (Supplementary Table 2), with the associations seen diminishing after further adjustment for BMD (Model 3: males – 1.84, 1.11-3.04, 0.02; females - 1.40, 0.92-2.13, 0.11).

### Mutual Adjustment

When each GM was mutually adjusted for the other two GMs, along with demographic characteristics and BMD, there was less evidence for an association with hip fracture in both combined and sex-stratified analysis. All results fell below the statistical significance threshold for multiple testing (Figure 5, Supplementary Table 2).

### Association between HSMs and hip fracture

Each HSM was initially assessed for its association with hip fracture. In the unadjusted combined-sex analysis, there was evidence of a strong positive association between HSM2 and hip fracture (Model 1: HR 1.32, 95% CI 1.11-1.56, *P*= 1.47×10^-3^) (Figure 4, Table 2). This association persisted upon adjustment for demographic characteristics and BMD (Model 3: 1.31, 1.11-1.55, 1.51×10^-3^). HSM2 captures features of a narrower FNW, a higher NSA, and reduced acetabular coverage (Figure 2). No other HSMs were found to be associated with hip fracture in combined-sex analysis.

**Table 2:** Cox proportional hazard results for the association between each hip shape mode and hip fracture in combined sex analysis.

|  | **Model 1** |  | **Model 2** |  | **Model 3** |  | **Model 4** |  |
| --- | --- | --- | --- | --- | --- | --- | --- | --- |
| **Exposure** | **HR [95% CI]** | **p-value** | **HR [95% CI]** | **p-value** | **HR [95% CI]** | **p-value** | **HR [95% CI]** | **p-value** |
| Hip shape mode 1 | 1.01 [0.85-1.20] | 0.91 | 0.94 [0.79-1.13] | 0.52 | 1.11 [0.92-1.33] | 0.27 | 1.12 [0.90-1.40] | 0.33 |
| Hip shape mode 2 | 1.32 [1.11-1.56] | $1.47 \times 10^{-3}$ | 1.36 [1.15-1.62] | $3.3 \times 10^{-4}$ | 1.31 [1.11-1.55] | $1.51 \times 10^{-3}$ | 1.30 [1.09-1.55] | $3.27 \times 10^{-3}$ |
| Hip shape mode 3 | 0.99 [0.84-1.18] | 0.94 | 1.13 [0.94-1.35] | 0.19 | 1.16 [0.97-1.39] | 0.10 | 1.10 [0.91-1.31] | 0.33 |
| Hip shape mode 4 | 0.87 [0.74-1.03] | 0.11 | 0.84 [0.71-1.00] | 0.05 | 0.88 [0.74-1.05] | 0.17 | 0.88 [0.71-1.07] | 0.20 |
| Hip shape mode 5 | 0.97 [0.82-1.15] | 0.74 | 0.98 [0.84-1.16] | 0.86 | 1.02 [0.87-1.21] | 0.79 | 0.99 [0.83-1.17] | 0.89 |
| Hip shape mode 6 | 1.09 [0.92-1.29] | 0.31 | 1.00 [0.84-1.19] | 1.00 | 0.96 [0.80-1.14] | 0.63 | 0.96 [0.79-1.16] | 0.68 |
| Hip shape mode 7 | 0.96 [0.81-1.14] | 0.66 | 0.99 [0.84-1.17] | 0.93 | 1.05 [0.88-1.24] | 0.60 | 1.11 [0.93-1.31] | 0.25 |
| Hip shape mode 8 | 1.00 [0.84-1.19] | 0.99 | 1.02 [0.86-1.21] | 0.82 | 0.95 [0.80-1.12] | 0.53 | 1.00 [0.83-1.21] | 0.98 |
| Hip shape mode 9 | 0.89 [0.75-1.06] | 0.19 | 0.96 [0.81-1.15] | 0.68 | 0.99 [0.82-1.18] | 0.87 | 0.90 [0.75-1.08] | 0.26 |
| Hip shape mode 10 | 0.99 [0.84-1.18] | 0.93 | 0.95 [0.80-1.13] | 0.59 | 0.97 [0.82-1.15] | 0.74 | 0.91 [0.76-1.09] | 0.30 |

Hazard ratios (HR) with 95% confidence intervals (CI) and p-values are shown for each hip shape mode and their association to hip fracture. Model 1 = unadjusted; model 2 = adjusted for age, sex, height, and weight; model 3 = adjusted for model 2 plus bone mineral density; model 4 = adjusted for model 3 plus the geometric measures.

In female sex-stratified analysis (Supplementary Table 1), HSM2 showed a positive association with hip fracture when adjusted for demographic characteristics (Model 2: 1.37, 1.11-1.68, 2.79×10^-3^). Apart from this, sex-stratified analyses failed to show statistical evidence for an association with hip fracture potentially because they were underpowered.

To evaluate the association between each HSM and hip fracture risk, independent of the hip shape components captured by GMs, each HSM was further adjusted for all three GMs (FNW, FHD, HAL) (Figure 4, Table 2). Analysis of all participants showed that the associations seen in Models 1, 2, and 3 were maintained after adjusting for demographic characteristics, BMD, and GMs. HSM2 emerged as the only HSM to show strong evidence of an association with hip fracture in this model (Model 4: 1.30, 1.09-1.55, 3.27×10^-3^). In sex-stratified analysis (Supplementary Table 1), none of the associations met the Bonferroni-adjusted p-value threshold. However, HSM2 showed weak evidence of an association with hip fracture when fully adjusted in both females and males (Model 4: females - 1.27, 1.03-1.57, 0.02; males - 1.34, 0.96-1.84, 0.06). Additionally, in males, HSM9 continued to show weak evidence of a negative association with hip fractures after full adjustment (Model 4: 0.66, 0.48-0.89, 0.01). No other HSM was associated with hip fracture when fully adjusted in either sex.

### Composite model

The composite model (Figure 3) showed that the overall at-risk shape, which is represented by the solid line, included a narrower FNW, reduced acetabular coverage, smaller greater trochanters, and a smaller FHD. This closely reflects HSM2, which shares these shape characteristics.

## Discussion

This large, longitudinal cohort study explored the relationship between DXA-derived HSMs and GMs with hip fracture risk. The findings indicate that HSM2, characterised by a narrower FNW, higher NSA, smaller femoral head, and reduced acetabular coverage, was positively associated with hip fracture risk, even after adjusting for age, sex, height, weight and BMD. While GMs (FNW, FHD, HAL) also showed associations with hip fracture when adjusted for the same covariates, these relationships attenuated upon mutual adjustment, confirming their inter-relatedness. In contrast, HSM2 retained its association with hip fracture after accounting for GMs, suggesting that HSM2 captures additional information beyond these three measures of hip geometry.

Currently, there are few comparative studies in the literature that have investigated the association between SSM-derived hip shape and hip fractures. Furthermore, these studies have derived their SSM from different populations, meaning it is not possible to draw direct comparisons between specific HSMs. For instance, Gregory et al. applied a SSM consisting of 29 points outlining the femoral head and neck to standard radiographs in a small group of females (26 cases, 24 controls)^9^. They found that SSM-derived hip shape predicted fracture risk after adjusting for height and weight. Specifically, a HSM with a longer, narrower femoral neck and a higher NSA was more likely to fracture, reflecting the at-risk shape identified in this study. However, their sample size was considerably smaller than that of our current study and the outline points on the radiographs used to perform SSM did not include the lesser trochanter. Baker-LePain et al. used a similar approach in a nested case-control study involving Caucasian females (168 cases, 231 controls)^8^. They employed a larger number of outline points (n=60) than Gregory et al. (n=29), and their model included the lesser trochanter. They found that hips exhibiting extreme values of HSM4, characterised by a narrower FNW, increased femoral neck length, and a smaller femoral head, were associated with hip fracture. These features closely resemble the at-risk hip shape identified in this study (narrower FNW and smaller femoral head). Although Baker-LePain et al. adjusted for age, body mass index, and femoral neck BMD, they, like Gregory et al., only included females within their analyses, leaving it unclear whether the observed relationships are sex-specific. Goodyear et al. performed SSM using 72 outline points on DXA scans of females aged over 75 years (182 subjects, 364 controls)^24^. The authors identified a hip shape associated with fracture that also closely resembles the findings of our study, including a narrower FNW, greater NSA, reduced acetabular coverage, and smaller greater trochanters. This study offers the closest comparison to the present analysis as it was performed on DXA scans and used similar outline points, including the acetabulum and lesser trochanter. However, the sample size was smaller, and analysis focused on females only. This limitation is significant because HSMs are known to be influenced by sex^25^, and our study found notable differences in HSMs between the sexes. Furthermore, none of the studies adjusted for GMs.

Although the at-risk hip shape (HSM2) identified in this study was characterised by a narrower FNW, the analysis of GMs and hip fracture revealed that a wider FNW was associated with hip fracture (Figure 5, Supplementary Table 2). This finding has been reported in other observational studies^26–28^, including a recent genetic analysis^29^ that found that individuals with a genetic predisposition to a greater FNW were at higher risk of fracture. The seemingly contradictory findings between HSM and GMs regarding FNW and fracture risk may be attributable to several factors. GMs objectively quantify individual aspects of hip morphology, meaning that bone size can impact the magnitude of the measurement. For example, larger individuals are likely to have a bigger femur across all dimensions; thus, a taller and heavier person would be expected to have a larger FNW and HAL. Moreover, FNW is highly correlated with height and moderately correlated with weight (Supplementary Figure 1), a relationship that has been consistently reported in other studies^11,30^, highlighting the significant influence of demographic characteristics, such as height and weight, on FNW. In contrast, SSM employs Procrustes analysis to align and scale hip outlines based on shape, effectively capturing bone morphology while excluding the influence of individual size. This ability to isolate shape from size is important because HSM2 remained associated with hip fracture risk, independent of FNW, FHD, and HAL. This suggests that these individual measures are not independently driving hip fracture risk. Instead, the interactions and combined influence of these factors, effectively captured by SSM, likely contribute to fracture risk. Ratios of GMs have been suggested as an alternative, aiming to reduce the influence of correlation by standardising measures against one GM^13^. However, SSM still outperformed ratio values in a previous small study^9^.

Previous research has explored sex differences in hip shape^23,25,31,32^; few studies have examined these differences within the context of hip fractures. HSM2 showed similar effect sizes between the sexes, but a notable difference was seen with HSM9 (Supplementary Table 1). Studies of individual hip shape measures have shown that females tend to have a smaller FHD, narrower FNW, and shorter femoral neck length compared to males^32^, which likely reflects that females are typically smaller than males. Similarly, Frysz et al. found sex differences in HSMs, with females exhibiting a narrower FNW and smaller lesser trochanter compared with males^25^. This finding is noteworthy, particularly given the weak evidence of a negative association with hip fracture seen with HSM9 in males (Supplementary Table 1). HSM9 was characterized by a larger lesser trochanter but a narrower femoral neck, suggesting that a larger lesser trochanter, a feature more common in male hip shapes, could offer some protective effect against hip fracture. Since the lesser trochanter serves as the insertion point for hip flexor muscles^33^ its size could be indicative of muscle mass. Given that sarcopenia (loss of muscle mass and function)^34,35^ is a known risk factor for hip fracture^36–39^, a larger lesser trochanter may represent a proxy for muscle strength and function, potentially reducing fracture risk in males. Additionally, innate female hip shape characteristics may predispose females to a higher fracture risk, as they often exhibit features linked to fractures, such as a narrower FNW. Interestingly, although HSM9 included a narrower FNW, similar to the fracture-prone HSM2, this reinforces the idea that fracture risk is influenced by multiple interacting shape constituents rather than any single measurement.

This study has several key strengths. Its large sample size and population-based design greatly enhances the representativeness of the findings, thereby improving the reliability of effect estimates. The study also simultaneously examined the relationship between SSM-derived hip shape and GMs with hip fracture, allowing for a direct comparison of these two methods and an evaluation of their independent associations with fracture risk. One of the limitations of SSM is that each study uses a different population to derive their HSMs, thus you cannot compare across models. This UK Biobank model could provide a reference for other populations. Both the SSM-derived hip shape and the GMs were semi-automatically derived from DXA scans, requiring minimal manual point correction. Given the widespread use of DXA scans in clinical practice for assessing osteoporosis, this approach makes accommodating SSM-derived hip shape measures through tools like FRAX a feasible option. Additionally, the inclusion of both combined and sex-stratified analyses represents a significant strength of this study. While many studies primarily examine post-menopausal females, this study also included male participants, providing valuable insights into male hip shape and its role in fracture risk.

There are limitations to this study. As an observational study, it cannot establish causality. Further research to understand the factors driving the association between HSM2 and hip fracture risk is needed, although a recent study using genetic data found evidence of a causal association between HSM2 and hip fracture in the same population^40^. NSA could not be derived from the DXA scans due to the limited view of the femoral shaft. Given that prior studies have linked higher NSA to hip fracture, and HSM2 represents a higher NSA, we were unable to determine if the association between HSM2 and hip fracture was independent of NSA^10,41^. The predominantly Caucasian study population may limit the generalisability of the findings. Notably, differences in hip shape have been reported between the UK Biobank cohort and the exclusively Chinese Shanghai Changfeng cohort^42^. The mean age of participants (63.7 years) may have reduced the study’s power, as hip fractures predominantly occur in older individuals^43^. However, as participants continue to be followed-up and additional DXA images from UKB become available, analysis can be repeated with more hip fracture cases, potentially strengthening findings. Since the analysis focused only on left hip DXA scans, and the side of the body the hip fracture occurred on is unknown, it is plausible that effect estimates could be biased towards the null. As a result, the true effect of hip shape on fracture risk may be underestimated or not fully captured in the analysis. Furthermore, using 2-dimensional DXA scans to assess the shape of a 3-dimentional structure may result in the loss of spatial detail; however, SSM can help mitigate these limitations by using proportional rather than absolute values of hip shape as described by GMs^9^.

In conclusion, this study examined SSM-derived hip shape using high-resolution DXA scans from a large cohort of UKB participants, demonstrating risk of incident hip fracture is higher with a narrower femoral neck, a higher femoral neck angle and reduced acetabular coverage. Importantly these associations were independent of a wide range of covariates including established measures of femoral geometry. Given that DXA scans are already routinely used to assess osteoporosis risk, it is conceivable that SSM-derived measures of hip shape could be accommodated into existing fracture risk tools such as FRAX^®^ to improve prediction. This approach could facilitate targeted preventative treatments for individuals with hip shapes resembling HSM2, thereby reducing the risk of hip fractures and alleviating the resultant morbidity and mortality.

## Data Availability

All data variables are available from UK Biobank. The BoneFinder® search model and the SSM can be requested via the BoneFinder® website for independent validation: https://bone-finder.com/

## ACKNOWLEDGEMENTS

This study has used the UK Biobank resource, access application 17295. The authors would like to thank Dr Monika Frysz, who was instrumental in deriving the hip shape modes in UK Biobank. For the purpose of Open Access, the author has applied a Creative Commons Attribution (CC BY) licence to any Author Accepted Manuscript version arising from this submission. NCH is supported by the UK Medical Research Council (MRC) [MC_PC_21003; MC_PC_21001] and National Institute for Health Research (NIHR) Southampton Biomedical Research Centre, University of Southampton, and University Hospital Southampton NHS Foundation Trust, UK.

## Author contributions

Contribution to study conception and design: SS, JHT, BGF, RAB

Contribution to acquisition of data: SS, AH, RE, FRS, JSG, RMA, CL, TC, NCH, JHT, RAB, BGF

Contribution to analysis and interpretation of data: SS, AH, RE, FRS, JSG, RMA, CL, TC, NCH, JHT, RAB, BGF

Drafting the article: SS, BGF, RAB

Reviewing the final manuscript: SS, AH, RE, FRS, JSG, RMA, CL, TC, NCH, JHT, RAB, BGF

## Conflict of interest statement

The authors have no conflicts of interest to disclose.

## Ethics statement

This study is overseen by the UKB Ethics Advisory Committee, and ethical approval was given by the National Information Governing Board for Health and Social Care and North-West Multi-centre Research Ethics committee (11/NW/0382). All participants provided informed consent for their data to be used in the study.

## Funding statement

SS and AH were self-funded undergraduate students. BGF is supported by an NIHR Academic Clinical Lectureship and an Academy of Medical Sciences Starter Grant (SGL030\1057). RB, RE, FS and MJ were supported by a Wellcome Trust collaborative award (209233/Z/17/Z). CL is funded by a Sir Henry Dale Fellowship jointly funded by the Wellcome Trust and the Royal Society (223267/Z/21/Z). For the purposes of open access, the authors have applied a CC BY public copyright licence to any Author Accepted Manuscript version arising from this submission.

## SUPPLEMENTARY TABLES AND FIGURES

**Supplementary Table 1:** Cox proportional hazard results for the association between each hip shape mode and hip fracture in sex-stratified analysis.

|  | **Model 1** |  | **Model 2** |  | **Model 3** |  | **Model 4** |  |
| --- | --- | --- | --- | --- | --- | --- | --- | --- |
| **Exposure** | **HR [95% CI]** | **p-value** | **HR [95% CI]** | **p-value** | **HR [95% CI]** | **p-value** | **HR [95% CI]** | **p-value** |
| **Males** |  |  |  |  |  |  |  |  |
| Hip shape mode 1 | 1.12 [0.83-1.52] | 0.47 | 1.09 [0.80-1.47] | 0.59 | 1.28 [0.94-1.75] | 0.12 | 1.32 [0.89-1.97] | 0.17 |
| Hip shape mode 2 | 1.43 [1.06-1.93] | 0.02 | 1.39 [1.03-1.88] | 0.03 | 1.38 [1.02-1.86] | 0.04 | 1.34 [0.98-1.84] | 0.06 |
| Hip shape mode 3 | 1.20 [0.88-1.62] | 0.25 | 1.25 [0.92-1.69] | 0.16 | 1.27 [0.94-1.73] | 0.12 | 1.17 [0.86-1.60] | 0.33 |
| Hip shape mode 4 | 0.79 [0.59-1.07] | 0.13 | 0.77 [0.57-1.04] | 0.09 | 0.79 [0.58-1.07] | 0.13 | 0.72 [0.50-1.03] | 0.08 |
| Hip shape mode 5 | 0.84 [0.62-1.15] | 0.28 | 0.85 [0.63-1.16] | 0.32 | 0.92 [0.67-1.26] | 0.59 | 0.87 [0.63-1.20] | 0.40 |
| Hip shape mode 6 | 1.00 [0.73-1.36] | 1.00 | 0.99 [0.73-1.35] | 0.97 | 0.96 [0.70-1.31] | 0.79 | 0.94 [0.68-1.31] | 0.73 |
| Hip shape mode 7 | 1.10 [0.82-1.47] | 0.53 | 1.14 [0.85-1.52] | 0.39 | 1.20 [0.89-1.61] | 0.24 | 1.29 [0.96-1.74] | 0.09 |
| Hip shape mode 8 | 0.98 [0.73-1.32] | 0.89 | 0.98 [0.73-1.31] | 0.89 | 0.90 [0.67-1.21] | 0.48 | 0.96 [0.70-1.32] | 0.81 |
| Hip shape mode 9 | 0.72 [0.54-0.98] | 0.04 | 0.70 [0.52-0.95] | 0.02 | 0.73 [0.54-0.99] | 0.04 | 0.66 [0.48-0.89] | 0.01 |
| Hip shape mode 10 | 0.95 [0.72-1.26] | 0.73 | 0.94 [0.71-1.25] | 0.68 | 0.97 [0.74-1.28] | 0.84 | 0.91 [0.67-1.23] | 0.53 |
| **Females** |  |  |  |  |  |  |  |  |
| Hip shape mode 1 | 0.83 [0.67-1.03] | 0.09 | 0.87 [0.70-1.09] | 0.22 | 1.02 [0.82-1.28] | 0.83 | 1.04 [0.80-1.36] | 0.76 |
| Hip shape mode 2 | 1.27 [1.04-1.56] | 0.02 | 1.37 [1.11-1.68] | $2.79 \times 10^{-3}$ | 1.26 [1.03-1.54] | 0.02 | 1.27 [1.03-1.57] | 0.02 |
| Hip shape mode 3 | 1.05 [0.84-1.31] | 0.68 | 1.07 [0.86-1.34] | 0.54 | 1.13 [0.90-1.40] | 0.29 | 1.08 [0.86-1.35] | 0.52 |
| Hip shape mode 4 | 0.97 [0.78-1.19] | 0.75 | 0.87 [0.71-1.08] | 0.21 | 0.94 [0.76-1.15] | 0.54 | 0.96 [0.75-1.23] | 0.76 |
| Hip shape mode 5 | 1.01 [0.83-1.24] | 0.89 | 1.05 [0.86-1.27] | 0.65 | 1.05 [0.86-1.29] | 0.60 | 1.03 [0.84-1.26] | 0.78 |
| Hip shape mode 6 | 1.03 [0.84-1.27] | 0.78 | 1.01 [0.82-1.24] | 0.96 | 0.96 [0.78-1.18] | 0.68 | 0.96 [0.76-1.22] | 0.75 |
| Hip shape mode 7 | 0.87 [0.71-1.07] | 0.18 | 0.94 [0.77-1.15] | 0.56 | 0.99 [0.81-1.22] | 0.96 | 1.04 [0.84-1.27] | 0.73 |
| Hip shape mode 8 | 1.05 [0.85-1.29] | 0.68 | 1.04 [0.85-1.28] | 0.70 | 0.98 [0.80-1.21] | 0.87 | 1.03 [0.82-1.29] | 0.82 |
| Hip shape mode 9 | 1.14 [0.91-1.41] | 0.25 | 1.15 [0.92-1.43] | 0.22 | 1.15 [0.92-1.44] | 0.21 | 1.07 [0.85-1.35] | 0.56 |
| Hip shape mode 10 | 1.01 [0.81-1.26] | 0.92 | 0.97 [0.78-1.21] | 0.80 | 0.96 [0.77-1.19] | 0.68 | 0.91 [0.72-1.14] | 0.40 |

Hazard ratios (HR) with 95% confidence intervals (CI) and p-values are shown for each hip shape mode and their association to hip fracture. Model 1 = unadjusted; model 2 = adjusted for age, height, and weight; model 3 = adjusted for model 2 plus bone mineral density; model 4 = adjusted for model 3 plus the geometric measures.

**Supplementary Table 2:** Cox proportional hazard results for the association between each geometric measurement and hip fracture.

|  | **Model 1** |  | **Model 2** |  | **Model 3** |  | **Model 4** |  |
| --- | --- | --- | --- | --- | --- | --- | --- | --- |
| **Exposure** | **HR [95% CI]** | **p-value** | **HR [95% CI]** | **p-value** | **HR [95% CI]** | **p-value** | **HR [95% CI]** | **p-value** |
| **Combined sex** |  |  |  |  |  |  |  |  |
| FNW (mm) | 1.15 [0.97-1.36] | 0.11 | 2.45 [1.80-3.33] | $1.17 \times 10^{-8}$ | 1.77 [1.30-2.43] | $3.26 \times 10^{-4}$ | 1.31 [0.88-1.96] | 0.19 |
| FHD (mm) | 1.12 [0.95-1.33] | 0.17 | 2.42 [1.76-3.32] | $5.20 \times 10^{-8}$ | 1.89 [1.39-2.57] | $4.48 \times 10^{-5}$ | 1.47 [0.96-2.25] | 0.07 |
| HAL (mm) | 1.08 [0.91-1.28] | 0.39 | 1.85 [1.34-2.55] | $1.93 \times 10^{-4}$ | 1.61 [1.18-2.21] | $3.08 \times 10^{-3}$ | 1.21 [0.84-1.74] | 0.31 |
| **Males** |  |  |  |  |  |  |  |  |
| FNW (mm) | 2.17 [1.44-3.25] | $1.99 \times 10^{-4}$ | 2.13 [1.33-3.43] | $1.74 \times 10^{-3}$ | 1.75 [1.08-2.82] | 0.02 | 1.17 [0.63-2.18] | 0.62 |
| FHD (mm) | 2.30 [1.54-3.44] | $4.50 \times 10^{-5}$ | 2.36 [1.45-3.84] | $5.54 \times 10^{-4}$ | 2.01 [1.28-3.14] | $2.26 \times 10^{-3}$ | 1.61 [0.85-3.06] | 0.14 |
| HAL (mm) | 2.07 [1.37-3.11] | $4.84 \times 10^{-4}$ | 2.01 [1.19-3.37] | 0.01 | 1.84 [1.11-3.04] | 0.02 | 1.39 [0.78-2.45] | 0.26 |
| **Females** |  |  |  |  |  |  |  |  |
| FNW (mm) | 2.88 [2.05-4.06] | $1.40 \times 10^{-9}$ | 2.71 [1.80-4.08] | $1.63 \times 10^{-6}$ | 1.70 [1.11-2.59] | 0.01 | 1.39 [0.82-2.35] | 0.22 |
| FHD (mm) | 2.43 [1.73-3.40] | $2.60 \times 10^{-7}$ | 2.46 [1.62-3.74] | $2.36 \times 10^{-5}$ | 1.70 [1.11-2.60] | 0.01 | 1.35 [0.77-2.36] | 0.29 |
| HAL (mm) | 2.05 [1.48-2.85] | $1.58 \times 10^{-5}$ | 1.73 [1.15-2.62] | 0.01 | 1.40 [0.92-2.13] | 0.11 | 1.09 [0.67-1.75] | 0.73 |

Hazard ratios (HR) with 95% confidence intervals (CI) and p-values are shown for each geometric measure and their association to hip fracture. Model 1 = unadjusted; model 2 = adjusted for age, sex, height, and weight (no sex adjustment in sex-stratified analysis); model 3 = adjusted for model 2 plus bone mineral density; model 4 = adjusted for model 3 plus the remaining 2 geometric measures.
FNW = femoral neck width, FHD = femoral head diameter, HAL = hip axis length

**Supplementary Figure 1:**
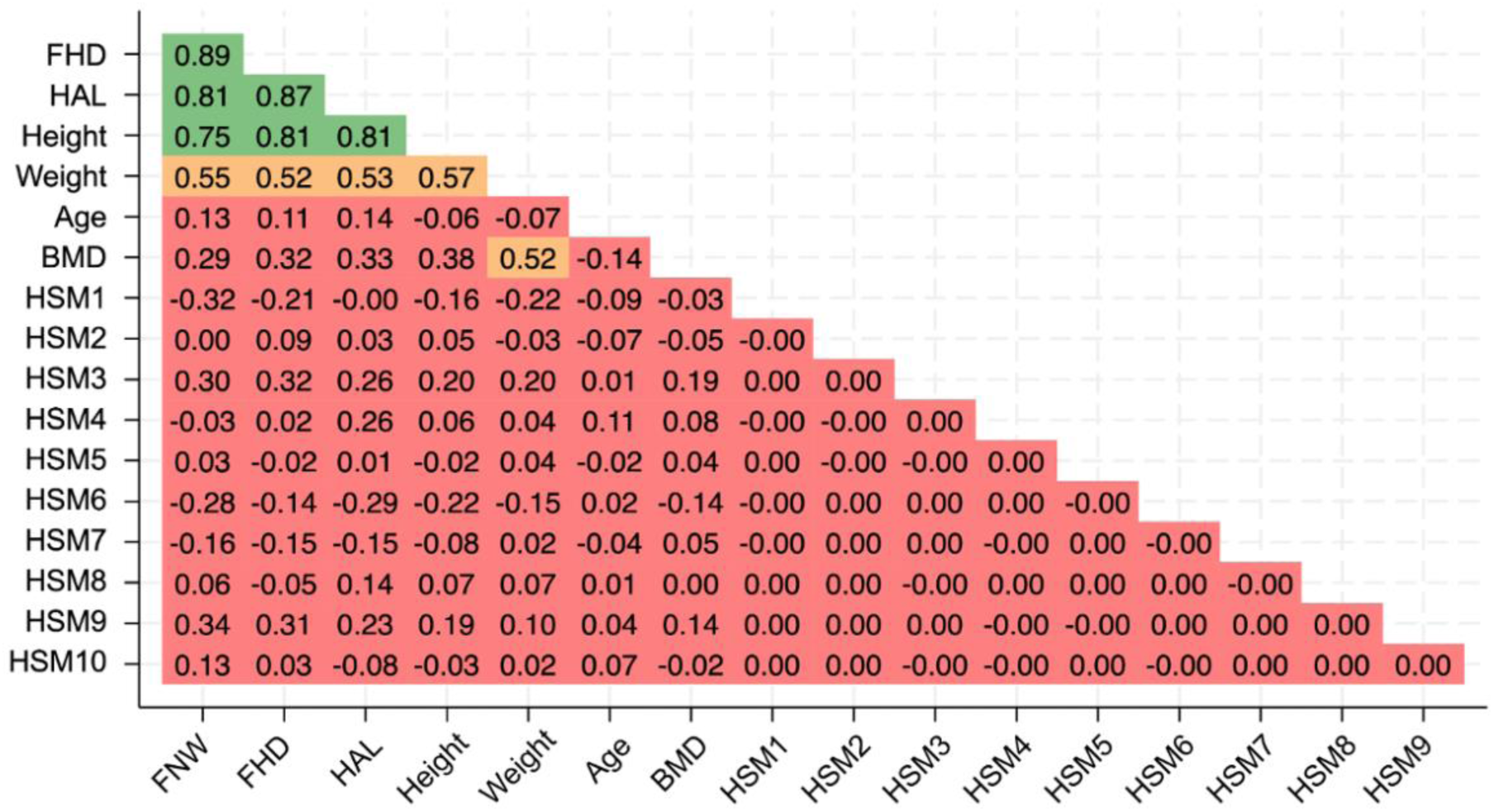
Pearson’s correlation matrix (r) showing the correlation between each HSM, GM (FHD, HAL, FNW), height, weight, age and BMD within the cohort. Green shows a strong correlation (r ≥0.7-1), orange shows a moderate correlation (r ≥0.5-<0.7), red shows a weak correlation (r <0.5). FHD = femoral head diameter, HAL = hip axis length, FNW = femoral neck width, BMD = bone mineral density, HSM = hip shape mode, GM = geometric measure.

## Notes

### Competing Interest Statement

The authors have declared no competing interest.

### Author Declarations

This study is overseen by the UK Biobank Ethics Advisory Committee. National Information Governing Board for Health and Social Care and North-West Multi-centre Research Ethics committee (11/NW/0382) gave ethical approval for this work. All participants provided informed consent for their data to be used in the study.

